# Gamified intervention to educate healthcare professionals on rational use of antimicrobials

**DOI:** 10.1101/2024.04.14.24305615

**Authors:** Sumana M Neelambike, Supreeta R Shettar, Yogeesh Maheshwarappa, G K Megha

## Abstract

**Background:** Antimicrobial resistance [AMR] is a global problem. It’s important to train health care professionals on rational use of antimicrobials to curb AMR.

**Methods:** This is a prospective interventional study conducted for the clinical practitioners, Undergraduates [MBBS/Interns], Post graduates and Pharmacy Students. A total of 50 participants were included in the study. The innovative games were administered for management of infections of all the different systems of the body in accordance with the ICMR treatment guidelines 2022 and latest IDSA guidelines involving different components. Pre-test and Post-test questionnaires were administered and evaluated.

**Results:** After the intervention, the knowledge on differentiating between bacterial and viral symptoms in respiratory tract infections and gastroenteritis improved from 48% to 94%. The practice of using right empirical choice of antimicrobials in the right dose for the right duration, based on the severity of the infection improved from 34 % to 82 %. The awareness/practice of using the right and rational combination of antibiotics improved from 44% to 84%. The knowledge on suspecting multi-drug resistant gram negative infections and other priority pathogens such as Methicillin-resistant *Staphylococcus aureus* [MRSA] and *Candida* infection improved from 32% to 78%. The practice of using certain antibiotics in specific infection sites based on their pharmacodynamics and pharmacokinetics improved from 20% to 76%. The knowledge on intrinsic resistance of certain microorganisms to specific antimicrobial agents improved from 15 % to 80%.

**Conclusion:** The gamified intervention successfully improved participants’ knowledge and awareness on rational antimicrobial use. The substantial improvements in all the aforementioned components highlight the positive impact of the intervention in promoting optimal antimicrobial use and curbing AMR. Innovative gamified interventions create better and long-lasting awareness ensuring the appropriate use of antimicrobials.

## Introduction

Antimicrobial resistance [AMR] has become a serious global health problem. The World Health Organization [WHO] has identified AMR as one of the top Ten public health concerns with the potential to kill 10 million people per year by 2050[1,2,3]. Initially, Penicillin’s discovery revolutionized the treatment of infectious diseases. Healthcare and global development were transformed to reduce mortality. Continued introduction of varied antimicrobials enhanced healthcare but their incorrect use at all levels of human, animal, and plant health unleashed AMR repercussions1.

In the face of rising AMR, LMIC’s health and development policies aiming to eliminate severe poverty [living on less than $1.90 per day] by 2030 have proven impossible. As per World Bank, the impact of AMR on healthcare expenses might vary from $300 billion to more than one trillion dollars by 20502. In May 2015, the World Health Assembly established a global action plan on antibiotic resistance. This includes strategic objectives aimed at raising awareness and knowledge of antimicrobial resistance, as well as optimizing the use of these medications. Continuous training is required both socially and professionally through various ways such as the use of media, conferences by specialists in the field, and Antimicrobial Optimization programs.

One of the most significant challenges in AMS Programs is the amount of knowledge students must master in a short period of time. AMR is a difficult concept to comprehend, and many [including pharmacists, and dentists] believe that the person develops resistance [9, 10]. Up-to-date knowledge of optimal antimicrobial use appears to be quite poor in medical students and practitioners. Medical educators face a significant challenge in developing and implementing instructional approaches that promise to increase students’ knowledge on antimicrobial resistance and antimicrobial stewardship.

Existing programs, frequently exhibit mixed results in terms of effectiveness. The conventional method of teaching students [lectures] has not actively engaged the students. It is largely a passive strategy that has not been proven to increase long-term memory or understanding of AMR concepts. It has been suggested that games actively include participants in the AMR learning process, which helps address these constraints [14, 15].

Games have recently been increasingly employed in AMS programs as games are more effective than conventional methods. Games can boost interest in a topic and reinforce previously provided information. They have been demonstrated to foster a competitive environment and boost student interaction. As a result, games could be viewed as a suitable supplement to existing healthcare initiatives, addressing the limitations of “conventional techniques” [16, 17]. The aim of the study is to find out how Games can increase the knowledge about antibiotic prescription practices by creating an engaging learning environment.

## Methods

This is a prospective interventional study conducted for the clinical practitioners, Undergraduate [MBBS/Interns], Post graduate and Pharmacy Students of JSS Medical College and JSS College of Pharmacy, Mysuru, South India. A total of 50 participants were included in the study. The participants were divided into 2 teams. The pre-test questionnaire was first administered to each participant for over a period of 20 minutes. The intervention was then conducted through the following:

1. Introduction about rational use of antibiotics, the importance of antimicrobial stewardship interventions in a tertiary hospital setting.
2. Playing innovative Games designed for management of infections of different systems [based on the latest ICMR Antimicrobial treatment guidelines 2022 and latest IDSA treatment guidelines].
3. Post-Test Questionnaire was administered to assess the effectiveness of intervention.

### Questionnaire

A multiple choice questionnaire covering all the systemic infections was designed for 150 marks with 30 questions, each carrying a weightage of 5 marks. The questions mainly focussed on the empirical choice of right antimicrobial agent in right dose and duration for a particular infection, antibiotics to be avoided in certain conditions based on pharmacokinetics and pharmacodynamics and intrinsic resistance. The participants answered the questionnaire independently for over a period of 20 minutes without using any reference materials, notes, or assistance. Thus the baseline knowledge was evaluated.

### Game Design

The innovative games were designed for management of infections of all the different systems of the body in accordance with the ICMR-AMR treatment guidelines 2022 and latest IDSA guidelines focussing on the following:

1. To differentiate between bacterial and viral features in common infections like respiratory tract infections and gastroenteritis as most often these infections are caused by the viruses yet they are treated with antibiotics.
2. To understand the right empirical choice of antimicrobials in right dose for right duration based on the severity of infection.
3. To use the right and rational combination of antibiotics.
4. To suspect the multi-drug resistant gram-negative infections and other priority pathogens causing infections like Methicillin resistant Staphylococcus aureus [MRSA] and Candida.
5. To highlight the antibiotics not useful in certain sites of infections based on pharmacodynamics and pharmacokinetic of the antibiotic.
6. Intrinsic resistance of some organisms to some antimicrobial agents
7. Miscellaneous-Dosage calculation based on renal functions, rational choice of investigations at different stages of infections, choosing the right antibiotic based in the Breakpoint Minimum inhibitory concentration Quotient [BMQ].

#### 1. Games to differentiate between bacterial/viral infections of respiratory tract and gastrointestinal tract

If in at least 30-50% of infections differentiation could be made, antibiotic treatment can be avoided which is a step forward in tackling AMR. For this basketball, bucketing the ball and monkeying with donkey games were used.

#### 2. To select the right antibiotic

Carom, Dart game, Hop scotch, where in Venn and Musical chair games were used.

#### 3. Combination of two antibiotics was taught by Death by double strike

#### 4. Selecting lower to higher antimicrobials based on severity of infections was taught through

a. Hierarchy by Severity b] Build your Bhurj Khalifa c] Where in Venn

#### 5. Selecting appropriate antimicrobials based on the site of infection was taught through

Drop the Dropout or Push to the Pandora games

#### 6. To understand the right dosage and duration of the right antibiotic

a. Fun with Numbers b] Bowling game and c] Statue game or “Pick your partner right” game was used.

##### 1. Respiratory tract infection

For understanding the rational use of antimicrobials of Respiratory tract infections, six games were played.

**Basketing the ball game**, to differentiate the features of viral and bacterial pharyngitis, sinusitis, and Lower respiratory tract infection [LRTI] such that unnecessary use of antibiotics could be avoided in at least 30-50% viral infections, a step forward in the fight against AMR.

**Fun with Numbers game**, to select the right dose of antibiotic and duration of treatment for bacterial Pharyngitis and LRTI.

**Bowling game**, to select the right antibiotics in the right dose and duration for bacterial sinusitis treatment.

**Death by double strike game**, to select the right set of antibiotics for CAP [Community-acquired Pneumonia] treatment.

**Hierarchy game** to select the right antibiotics for LRTI treatment based on the severity of the infection.

**Drop the dropout to Pandora** to understand the antibiotics to be avoided in respiratory tract infections during pregnancy with the help of a case scenario.

**Dart game** to understand the right antibiotics to be used and antibiotics to be avoided in LRTI.

##### 2. Gastrointestinal and Intra-abdominal infections

For understanding the rational use in gastro-enteritis, 2 games were administered.

a. **Monkeying with Donkey Game** to segregate the viral and bacterial features of gastroenteritis in different clinical scenarios such that unnecessary use of antibiotics could be prevented.
b. **Statue game** to select the right antibiotic with the right dose and duration for the treatment of bacillary dysentery, Cholera, Amoebiasis, and Giardiasis. For intra-abdominal infections.

a. **Build your Bhurj Khalifa game** to select the antibiotics for the treatment of mild, moderate, and severe Community-acquired intra-abdominal infections and Hospital-acquired intra-abdominal infections from lower to higher antimicrobials. This could prevent the use of higher antimicrobials for a milder infection and lower antibiotic for severe infections such that the outcome in the patient is better and cost effective. This can also slow down the evolving
b. **Paint your imagination game** to know a host of antimicrobials used to treat different intra-abdominal infections like liver abscess, cholecystitis, pancreatic abscess/necrosis with or without risk of candidiasis, etc.

##### 3. Urinary tract infections

In Urinary tract infections, three games were played.

a. Picture puzzle for treatment of uncomplicated cystitis
b. **Pneumonic** created for treatment options of Pyelonephritis [**AIM-PE]**, Prostatitis [**P-TIMES]**, and Epididymo-orchitis [**COLD]**
c. **Drop the Dropout to the Pandora game** to know the antibiotics to be avoided in the UTIs.
d. Fun with numbers game was used to understand the dosage and duration of empirical antibiotics used for uncomplicated cystitis and pyelonephritis.

##### 4. Acute Undifferentiated Fever

a. **Island and Woodland game** to understand the clinical features supporting the AUF.
b. **The Loop game** to know the causative agents of Fever with Jaundice, Fever with Sore throat, and Fever with Rash in AUF.
c. **Where in Venn Game**: This helps to select the right antibiotic for Enteric fever, Rickettsia fever, and Leptospirosis.
d. **Drop the Dropout** to the pandora helps to rationally choose laboratory investigation in AUF.

##### 5. Sepsis

Sepsis and Septic shock were taught by using the **pneumonic** and treatment of sepsis by using the **tall and short doctors in a race story**. Suspecting MDRO was taught through picture puzzles. MRSA suspicion and its treatment are taught by **story creation game**. Suspicion of Pseudomonas and Candida infections were taught through **pneumonic**.

##### 6. Skin and Soft Tissue Infection [SST]

**The dart game wa**s used to know the antibiotics that can reduce toxin production.

**Scrabble game** was used to teach the treatment options of SST.

**Bowling game**: Antibiotics that do not act on anaerobes and antibiotics having anti-tubercular activity taught through this.

**Colour your imagination** was played to learn the treatment of Necrotising fasciitis.

**Picture puzzles** and **where in Venn game** were used to treat deep neck space infection.

##### 7. Central Nervous System Infection

The story was used to teach pointers to the diagnosis of Acute Febrile Encephalopathy/ Acute encephalitis syndrome.

**Where in Venn** game was used to teach treatment of meningitis

## Results

The different components of rational use of antimicrobials assessed pre-test and post-test through innovative gamified interventions were:

I. To differentiate between bacterial and viral symptoms in common infections like respiratory tract infections and gastroenteritis, while most often these infections are caused by viruses yet they are treated with antibiotics.
II. To understand the right empirical choice of antimicrobials in the right dose for the right duration, based on the severity of the infection.
III. To use the right and rational combination of antibiotics.
IV. To suspect the Multi-drug resistant [MDR] gram-negative infections and other priority pathogens causing infections like Methicillin-resistant Staphylococcus aureus [MRSA] and Candida.
V. To highlight the antimicrobials not useful in certain sites of infections based on the pharmacodynamics and pharmacokinetics of the antimicrobials
VI. Intrinsic resistance of some organisms to some antimicrobial agents.
VII. Miscellaneous

Table 1 depicts the comparison between pre and post-test results of six different components on rational use of antibiotics

**Table 1.** depicts the comparison between pre and post-test results of six different components on rational use of antibiotics.

| Component | Pre-Test in % [Out of 50 participants] | Post-Test in % [Out of 50 participants] | p-value |
| --- | --- | --- | --- |
| I | 48 | 94 | < 0.05 |
| II | 34 | 82 | < 0.05 |
| III | 44 | 84 | < 0.05 |
| IV | 32 | 78 | < 0.05 |
| V | 20 | 76 | < 0.05 |
| VI | 15 | 80 | < 0.05 |

### Pre-Test result analysis

The knowledge on intrinsic resistance of certain microorganisms to specific antimicrobial agents [Component VI] was least during the pre-test [15 %]. The knowledge on suspecting multi-drug resistant gram negative infections and other priority pathogens such as Methicillin-resistant *Staphylococcus aureus* [MRSA] and *Candida* [Component IV] was also very low [32%]. The practice of using right empirical choice of antibiotics for the right duration based on the severity of the infection [Component II] was satisfactory [34%]. The practice of using certain antibiotics in specific infection sites based on their pharmacodynamics and pharmacokinetics [Component V] was better when compared to the other five components [20%]. The awareness/practice of using the right and rational combination of antibiotics [Component III] was also better when compared to the other five components [44%]. The knowledge about differentiating between bacterial and viral symptoms in common infections like respiratory tract infections and gastroenteritis [Component I] was the best when compared to the other five components [48%].

After the intervention, the knowledge about differentiating between bacterial and viral symptoms in common infections like respiratory tract infections and gastroenteritis improved from 48% to 94%. The practice of using right empirical choice of antimicrobials in the right dose for the right duration, based on the severity of the infection improved from 34 % to 82 %. The awareness/practice of using the right and rational combination of antibiotics improved from 44% to 84%. The knowledge on suspecting multi-drug resistant gram negative infections and other priority pathogens such as Methicillin-resistant *Staphylococcus aureus* [MRSA] and *Candida* infection improved from 32% to 78%. The practice of using certain antibiotics in specific infection sites based on their pharmacodynamics and pharmacokinetics improved from 20% to 76%. The knowledge on intrinsic resistance of certain microorganisms to specific antimicrobial agents improved from 15 % to 80%.

## Discussion

The study’s findings show that the gamified intervention was effective in improving the knowledge and awareness on the rational use of antimicrobials involving different components. The significant improvements observed in all six components indicate that the intervention had a positive impact on promoting rational and responsible antimicrobial use.

In this study, participants showed an improved ability to distinguish between bacterial and viral symptoms in common infections from 48% to 94% [Component I]. In the study done by Azevedo et al they have found that the knowledge of correct use of antibiotics for bacterial diseases rather than viral rose from 43% to 76% after the teaching activity [Azevedo et al., 2013]. The improvement indicates that the intervention assisted the participants in distinguishing between bacterial and viral aetiologies, aiding them in rational use of antimicrobials and avoiding the unnecessary use of antibiotics which is a way forward in curbing growing AMR.

Understanding the empirical selection of antimicrobials based on the severity of the infection [Component II] is critical for effective treatment. The significant improvement in this component indicates that participants gained a better understanding of choosing the appropriate antimicrobial agents in appropriate dose, and treatment duration based on the severity and site of the infection.

The significant improvement in the third component i. e awareness of using the correct and rational combination of antibiotics demonstrates the intervention’s impact on promoting knowledge on the antibiotic choice and combination, reducing the likelihood of inappropriate and ineffective antimicrobial use minimizing resistance development.

Component IV, detecting multi-drug resistant gram-negative infections and other priority pathogens is critical for appropriate treatment and implementation of infection control practices. Significant improvement in this component suggests that participants became more proficient at identifying these challenging infections, allowing for targeted interventions and improved patient outcomes.

Understanding the limitations of specific antibiotics at specific infection sites [Component V] is critical for avoiding unnecessary antibiotic use. The significant improvement in this component suggests that participants learnt about the pharmacodynamics and pharmacokinetics of antibiotics, allowing them to take better informed decisions about their appropriate use.

The intrinsic resistance of some organisms to specific antimicrobial agents [Component VI] is useful for tailoring treatment approaches. The significant improvement in the knowledge on this component helps the participants to avoid use of irrational antimicrobials for certain organisms causing infections.

## Conclusion

The gamified interventions employed in this study successfully improved participants knowledge and awareness on rational use of antimicrobials. These improvements are a step ahead in curbing AMR, and optimizing patient outcomes by reducing unnecessary antimicrobial prescriptions. Continued efforts to enhance the knowledge and create awareness on rational use of antimicrobials is the need of the hour for combating global challenge of AMR and preserve the efficacy of the antimicrobial agents for the future generations. Innovative gamified interventions create better and long-lasting awareness promoting rational use of antimicrobials to curb the growing AMR.

## Data Availability

All data produced in the present work are contained in the manuscript

## Author Contributions

SMN identified the need of the education, devised the innovative games on awareness and designed the questionnaire and the study. She also got involved in conducting the study, analysis of the report compiled and contributed to prepare the manuscript. SRS and YM prepared the material required for the games.

## Data availability statement

No additional data available.

## Ethical clearance

Name of Ethical Committee: Institutional Ethical Committee, JSS Medical College, JSSAHER. This study involves human participants but an Ethics Committee(s) or Institutional Board(s) exempted this study because the study did not involve any diagnostic or therapeutic intervention program involving any human subjects.

